# Modelling the Impact of Delaying Vaccination Against SARS-CoV-2 Assuming Unlimited Vaccines Supply

**DOI:** 10.1101/2021.02.22.21252189

**Authors:** Marcos Amaku, Dimas Tadeu Covas, Francisco Antonio Bezerra Coutinho, Raymundo Soares Azevedo, Eduardo Massad

## Abstract

**Background:** At the moment we have more than 109 million cases and 2.4 million deaths around the world and vaccination represents the only hope to control the pandemic. Imperfections in planning vaccine acquisition and difficulties in implementing distribution among the population, however, have hampered the control of the virus so far.

**Methods:** We propose a new mathematical model to estimate the impact of vaccination delay against COVID-19 on the number of cases and deaths by the disease in Brazil. We apply the model to Brazil as a whole and to the State of Sao Paulo, the most affected by COVID-19 in Brazil. We simulated the model for the populations of the State of Sao Paulo and Brazil as a whole, varying the scenarios related to vaccine efficacy and compliance from the populations.

**Results:** The model projects that, in the absence of vaccination, almost 170 thousand deaths and more than 350 thousand deaths until the end of 2021 for Sao Paulo and Brazil, respectively. If in contrast, Sao Paulo and Brazil had enough vaccine supply and so started a vaccination campaign in January with the maximum vaccination rate, compliance and efficacy, they could have averted more than 112 thousand deaths and 127 thousand deaths, respectively. In addition, that for each month of delay the number of deaths increases monotonically in a logarithm fashion, for both the State of Sao Paulo and Brazil as a whole.

**Conclusions:** Our model shows that the current delay in the vaccination schedules that is observed in many countries has serious consequences in terms of mortality by the disease and should serve as an alert to health authorities to speed the process up such that the highest number of people to be immunized is reached in the shortest period of time.

## Introduction

As the world struggles to implement vaccination schemes against SARS-CoV-2, limited production of doses, imperfections in planning vaccine acquisition and difficulties in implementing distribution among the population, have hampered the control of the virus so far [1]. As of 18 February 2021, 188 million people have been vaccinated around the world, which represents less than 3% of the total. In Brazil, the total number of vaccinated people so far is, around 2.5% of the target population [2]. The world vaccination rate currently is less than 4 million doses per day, a very timid rate [3]. Soit is no surprise that vaccination by itself has had so far little effect on the number of cases and deaths that continues to soar in many countries. At the moment we have more than 109 million cases and 2.4 million deaths around the world [4].

Although previous pandemics have demonstrated that pharmaceutical interventions are less important than non-pharmaceutical intervention in controlling the infection, there is a growing body of evidences that this will not be the case with the vaccines against COVID-19 [5-8].

Immediately after the emergence of SARS-CoV-2 in China, many laboratories around the world started the development of more than 100 types of different vaccines, short-circuiting in less than one year the usual time frame of new vaccines development and testing, which normally would be around ten years, a remarkable tour of force [6], [9].

There is a wide range of covid-19 vaccines being developed [10]. As of February 2021, there are 80 vaccine candidates in 212 clinical trials and 11 vaccines approved by at least one country [10].Of these, 80 vaccines are in the pipeline, of which 20 are in Phase 3 of clinical trials (four have already completed this phase) and 37in Phase 2 [10]. In the US, three vaccines completed Phase 3 trials, namely, Moderna, Pfizer, and Oxford-AstraZeneca, and two are still in Phase 3 [10].

However, in order to have significant impact on the course of the pandemic, safe and effective vaccines have to emerge in less time that it would take the affected populations to reach natural herd immunity because to wait to have herd natural immunity would be disastrous. Therefore, an unprecedented time-schedule to roll out any effective vaccine is urgently needed. Nevertheless, in many countries the vaccination is limited to certain individual groups and the distribution of enough doses for these individuals is very slow. We have at the moment only 188 million doses applied in 64 countries around the world [11].

Brazil has accumulated almost 10 million cases and more than 240 thousand deaths at the time of writing (18 February 2021) [12]. The state of Sao Paulo, the most populous in Brazil reported almost 2 million cases and 57 thousand deaths so far [13]. Notwithstanding the fact that Brazil is the third country with the highest number of cases and second with the highest number of deaths in the world, only two vaccines, Coronavac and Oxford-AstraZeneca have been licensed. Currently, just above 2% of the target population have received a first dose of one or the other vaccines [14].

This paper proposes a new model to estimate the impact of vaccination delay against Covid-19 on the number of cases and deaths by the disease in Brazil. We apply the model to Brazil as a whole and to the State of Sao Paulo, the most affected by COVID-19 in Brazil. This work is a theoretical exercise because it assumes that at every time of the pandemic there is enough vaccine, which is not realistic for the majority of countries.

### The model

The model is an extension of the one presented in [15] and has the following variables:

1. *Susceptible* individuals, denoted *S*(*t*), which can either be vaccinated with rate ν, or acquire the infection with rate β. Susceptible are born with rate Λ and die by other causes with rate μ;
2. *Vaccinated* individuals, denoted *V*(*t*), which are transferred from the susceptible state with the vaccination rate ν. The vaccine is assumed to have efficacy *q* and a fraction *w* of the susceptible complies with the vaccination policy. Vaccinated individuals die by other causes with rate μ;
3. *Failure* to be immunized individuals, denoted *FV*(*t*). A fraction (1-*q*) of those vaccinated susceptible fail to be immunized, and can either acquire the infection with the same rate β as those non-vaccinated susceptible or die by other causes with rate μ;
4. *Exposed* individuals, denoted *E*(*t*), are those individuals who acquired the infection but are still in the stage that precedes either the overtly diseased patients or the asymptomatic individuals (see below). Exposed individuals can either progress to an asymptomatic stage with rate δ_A_, or to full-blown COVID-19 patients with rate δ_I_ or die by other causes with rate μ. A fraction *p*_E_ of those exposed are infective to susceptibles;
5. *Asymptomatic* (or pauci-symptomatic) individuals, denoted *A*(*t*), who progressed from the exposed and are, therefore, infected with SARS-CoV-2 but show no or very few symptoms. Asymptomatic individuals can either die by natural causes or by the infection, with rates μ and α_A_, respectively, or recover from the infection with rate γ_A_. A fraction *p*_A_of these asymptomatic individuals are infective to susceptible;
6. *Infective* individuals, denoted *I*(*t*), are those individuals infected with SARS-CoV-2 and who show the characteristic clinical signs and symptoms of COVID-19. Infective individuals can either die by natural causes or by the infection, with rates μ and α_*I*_, respectively, or recover from the infection with rate γ_I_, or progress to hospitalized (*H*(*t*))or critically ill stages (*G*(*t*)) (see below) with rates σ_H_ and σ_G_, respectively;
7. *Hospitalized* individuals, denoted *H*(*t*), are individuals with full-blown COVID-19 but who do not require Intensive Care Unit support. These individuals can die by natural causes or by the infection, with rates μ and α_H_, respectively, or recover from the infection with rate γ_H_;
8. *Grave* patients, denoted *G*(*t*), are seriously ill patients requiring Intensive Care respiratory support. These individuals can die by natural causes or by the infection, with rates μ and α_H_, respectively, or recover from the infection with rate γ_G_; and finally
9. *Recovered* individuals, denoted *R*(*t*), are those individuals who recovered from the infection. They can die by natural causes with rate μ.

Figure 1 shows a diagram with the model’s stages and transitions.

**Figure 1.**
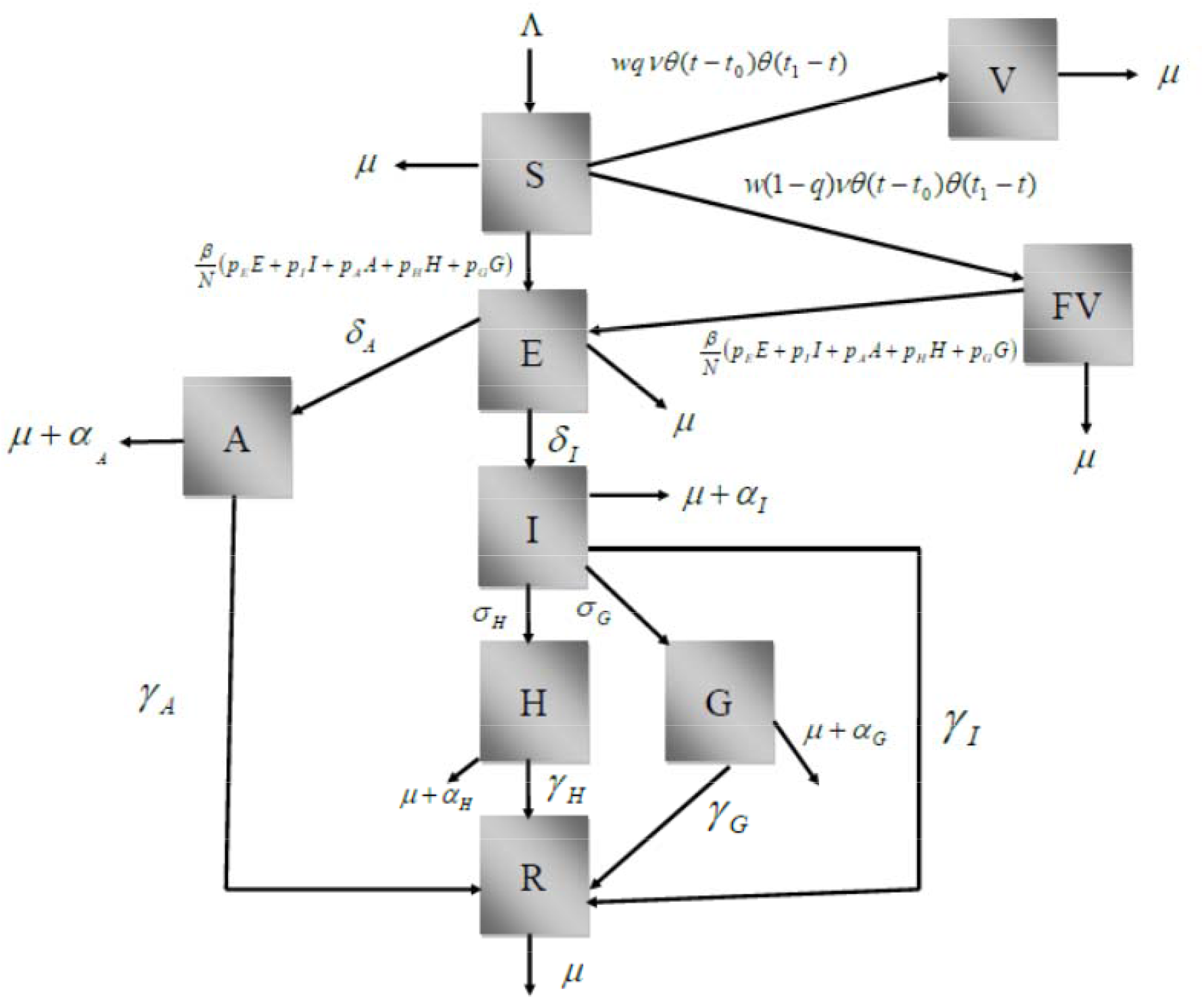
Diagram showing the model’s stages and transitions.

The model is described by the following set of equations:

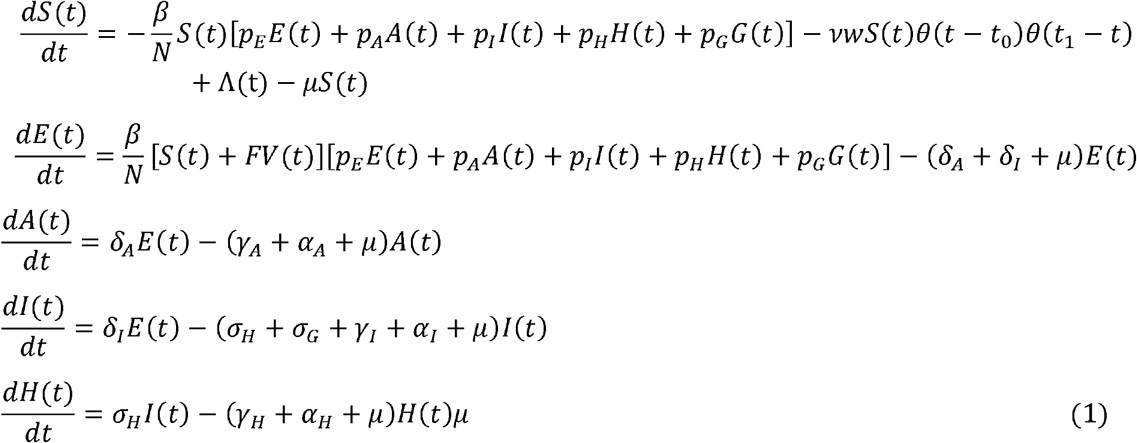

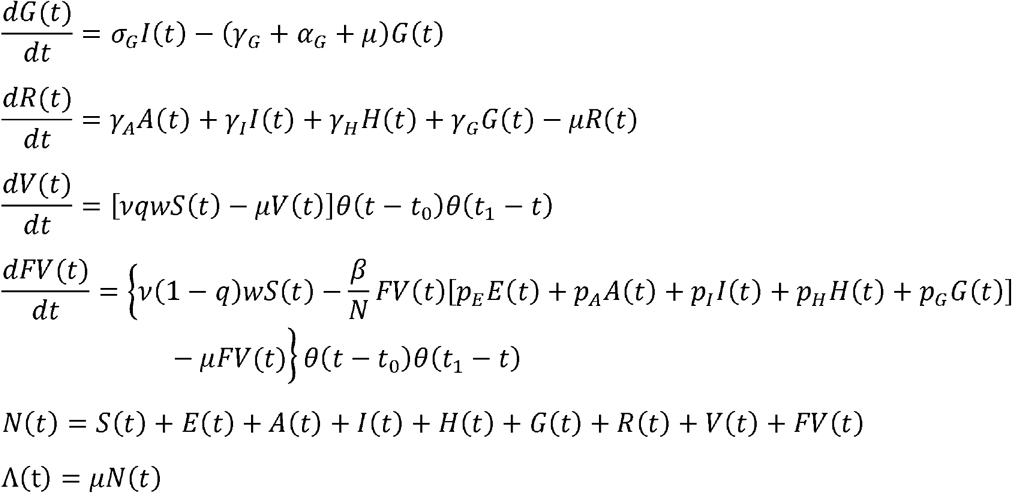

The incidence, *Inc*(*t*), is given by:

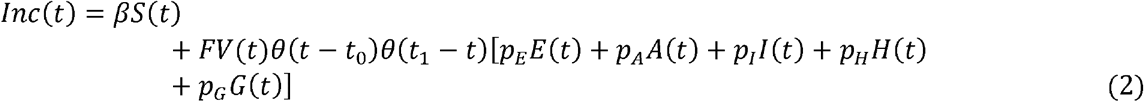

The total number of cases, *Cases*, is given by:

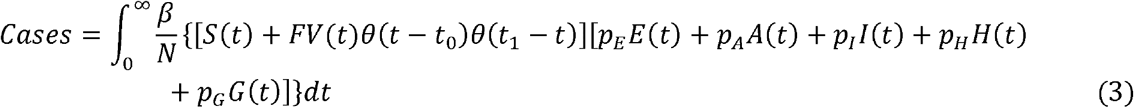

The total number of deaths, *Deaths* due to COVID-19,is given by:

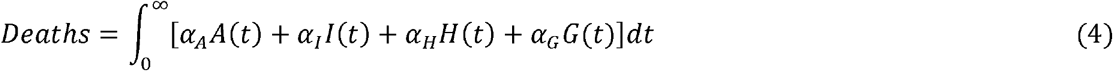

The total number of vaccinated individuals, *Vaccinated*, is given by:

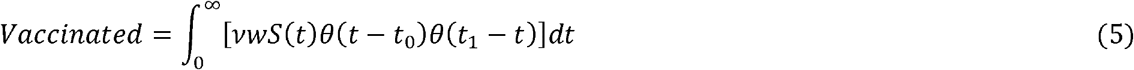

In table 1 we show the parameters used for the simulation of model (1) for Brazil and the State of São Paulo.

**Table 1.** Parameters used in the model for the State of São Paulo and Brazil.

| Parameter | Description | Value |  |
| --- | --- | --- | --- |
|  |  | São Paulo | Brazil |
| $\beta(t)$ | Potentially infective contact rate | Fitted (changes over time) | |
| $p_E$ | Infectivity of exposed individuals | 0.4* | 0.4* |
| $p_I$ | Infectivity of symptomatic individuals | 1.0* | 1.0* |
| $p_A$ | Infectivity of asymptomatic individuals | 1/3* | 1/3* |
| $p_H$ | Infectivity of hospitalized individuals | 0.01* | 0.01* |
| $p_G$ | Infectivity of ICU patients | 0.01* | 0.01* |
| $\mu$ | Natural mortality rate (life expectancy of 70 years) | $3.91 \times 10^{-5} \text{ days}^{-1}$ * | $3.91 \times 10^{-5} \text{ days}^{-1}$ * |
| $\delta_I$ | Rate of evolution from exposed to infected | $1/2 \text{ day}^{-1}$ * | $1/2 \text{ day}^{-1}$ * |
| $\delta_A$ | Rate of evolution from exposed to asymptomatic | $0.874 \text{ day}^{-1}$ ** | $0.366 \text{ day}^{-1}$ ** |
| $\gamma_I$ | Rate of recovery from infected | $1/3 \text{ day}^{-1}$ * | $1/3 \text{ day}^{-1}$ * |
| $\gamma_A$ | Rate of recovery from asymptomatic | $1/14 \text{ day}^{-1}$ * | $1/14 \text{ day}^{-1}$ * |
| $\gamma_H$ | Rate of recovery from hospitalized | $1/10 \text{ day}^{-1}$ * | $1/10 \text{ day}^{-1}$ * |
| $\gamma_G$ | Rate of recovery from ICU | $0.05 \text{ day}^{-1}$ ** | $0.0556 \text{ day}^{-1}$ ** |
| $\alpha_I$ | Disease-induced mortality rate for infected individuals | $5 \times 10^{-4} \text{ day}^{-1}$ * | $3 \times 10^{-4} \text{ day}^{-1}$ * |
| $\alpha_A$ | Disease-induced mortality rate for asymptomatic individuals | 0 * | 0 * |
| $\alpha_H$ | Disease-induced mortality rate for hospitalized individuals | $10^{-4} \text{ day}^{-1}$ ** | $5.56 \times 10^{-4} \text{ day}^{-1}$ ** |
| $\alpha_G$ | Disease-induced mortality rate for ICU patients | Fitted (changes over time) | |
| $\sigma_H$ | Hospitalization rate | $0.12 \text{ day}^{-1}$ ** | $0.0518 \text{ day}^{-1}$ ** |
| $\sigma_G$ | ICU admission rate | Fitted (changes over time) | |
| $v$ | Vaccination rate | Variable (from $5 \times 10^{-3} \text{ days}^{-1}$ to $10^{-1} \text{ days}^{-1}$) | |
| $q$ | Vaccination efficacy | Variable | |
| $w$ | Adherence to the vaccination campaign | Variable | |
| $K(t)$ | Notification ratio | Fitted (changes over time) | |
| $\Lambda(t)$ | Birth rate | Changes over time | |

\*assumed; \*\*fitted

In figure 2 we show the time that it would take to reach herd immunity (70% of the population) as a function of the vaccination rate, ν. This is calculated, approximately (neglecting mortality) as:

**Figure 2.**
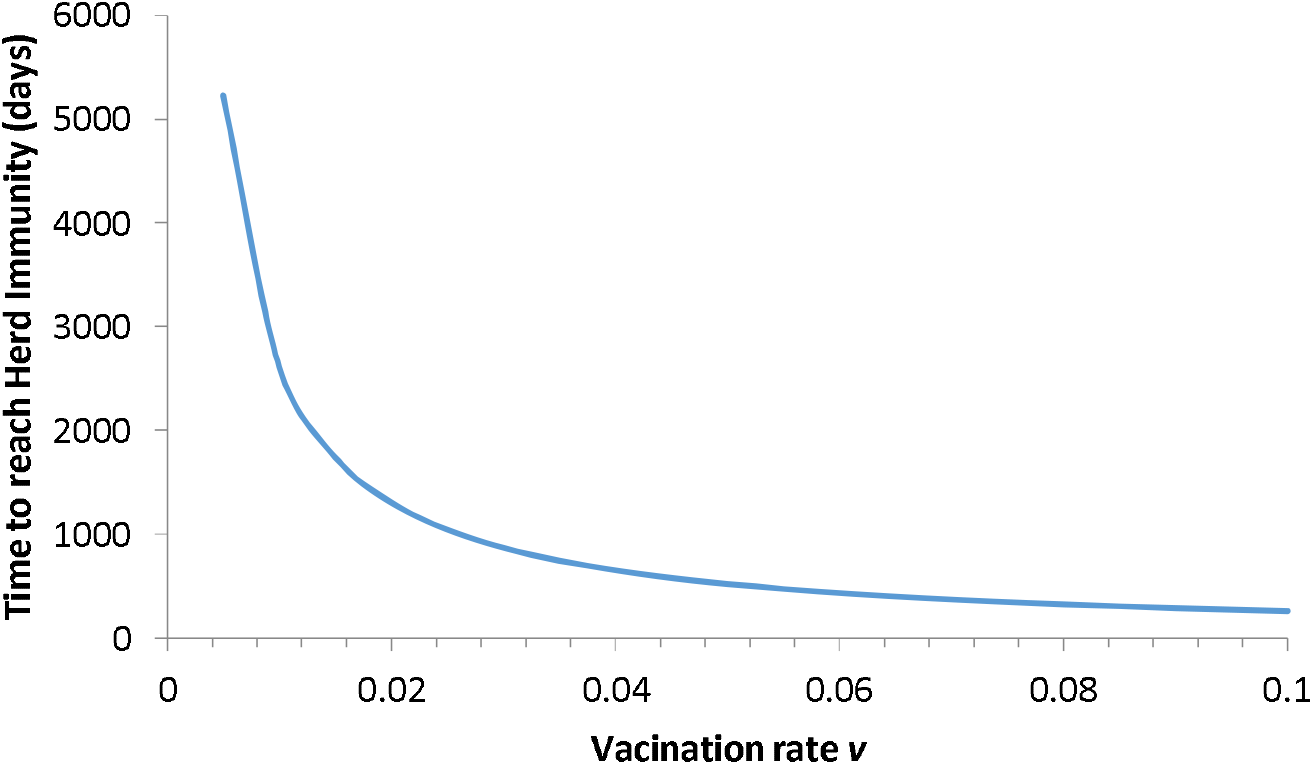
**Time in days taken to reach herd immunity as a function of vaccination rates as used in the simulation of the model.**

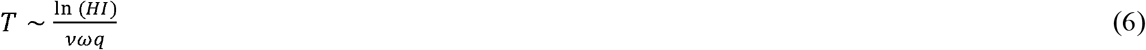

In equation (6), *T* means time to reach herd immunity (*HI*).

## Results

We simulated model (1) with the parameters as in table 1 for the two populations of the State of Sao Paulo and Brazil as a whole, varying the scenarios related to vaccine efficacy and compliance from the populations. We simulated efficacy with values of 50%, 70% and 90% and compliance with values of 50%, 70% and 80%. As mentioned above, we simulated the scenarios for Brazil as a whole and for the State of São Paulo. As shown in table 1, we have chosen values of vaccination rates that varied from 0.005 days^-1^ until 0.1 days^-1^. This implies that the time taken to reach herd immunity varied from 16 years to about 9 months, as shown in figure 2.

Below we show the results of the numerical simulations of the model.

### 1) Brazil

We fitted the model parameters simultaneously to the data of cumulative number of reported cases and deaths (figure 3) for Brazil until December 18, 2020. The fitting procedure is described in [16],[17].To estimate a 95% probability interval (shaded area in figure 3), we assumed a normal distribution for the contact rate with a standard deviation of 1.0%.

**Figure 3.**
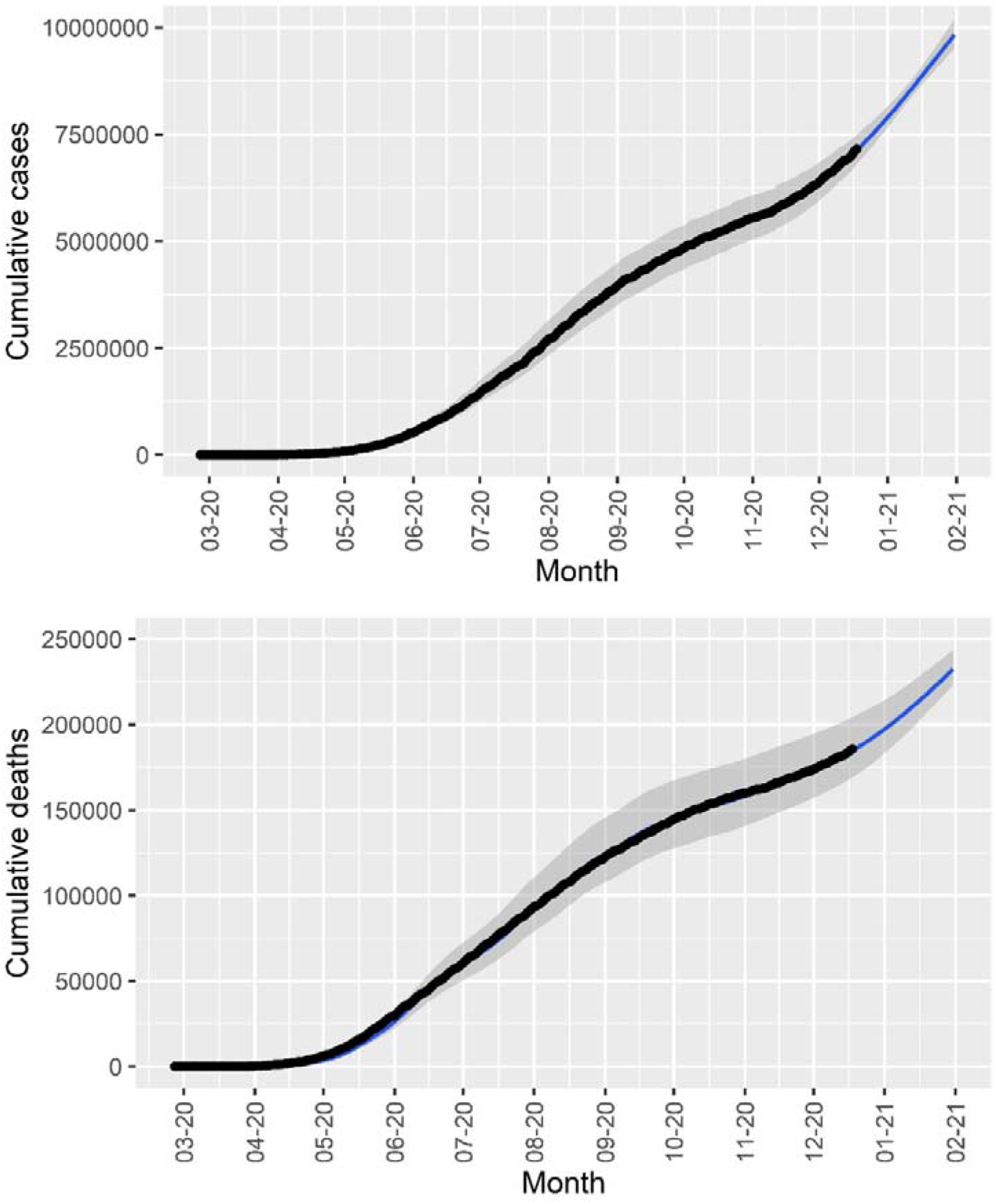
**Cumulative number of reported cases and deaths in Brazil (black dots) and the corresponding fitted model (blue lines). The solid lines and shaded area correspond, respectively, to median values and 95% probability intervals.**

In figure 4 we show the percentage of averted deaths until 31^st^ December 2021 for several scenarios simulated and for mass vaccination starting on January 21^st^, February 21^st^, March 21^st^, April 21^st^ and May 21^st^, for 3 combinations population compliance to the campaign and vaccine efficacy.

**Figure 4.**
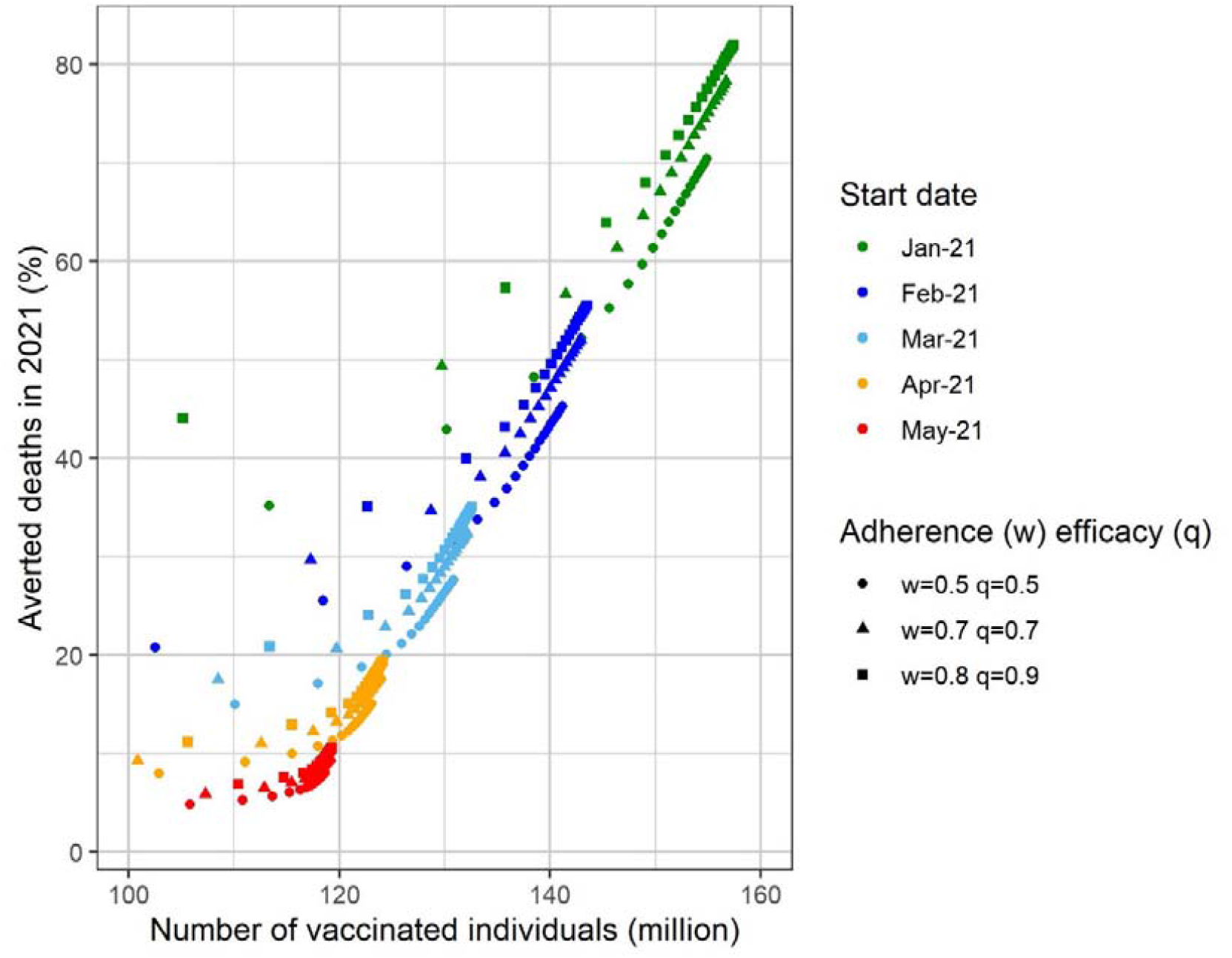
**Percentage of averted deaths in 2021 in Brazil as a function of the number of vaccinated individuals for different start dates for the vaccination campaign. Three different combinations of vaccination adherence (w) and vaccine efficacy (q) were considered: w=0.8 and q=0.9 (best-case scenario), w=0.7 and q=0.7 (baseline scenario) and w=0.5 and q=0.5 (worst-case scenario).**

It can be noted from figure 4 that if Brazil had started a mass vaccination campaign on January 21^st^ with the maximum compliance of 80%, a vaccine 90% efficacious, and a high vaccination rate (around 0.1 days^-1^), 80% of the expected deaths until December 31^st^ would be averted.

This result can also be seen in Figure 5 in which we show the percentage of averted deaths until the end of the year as a function of the vaccination rate for vaccination starting from January until May.

**Figure 5.**
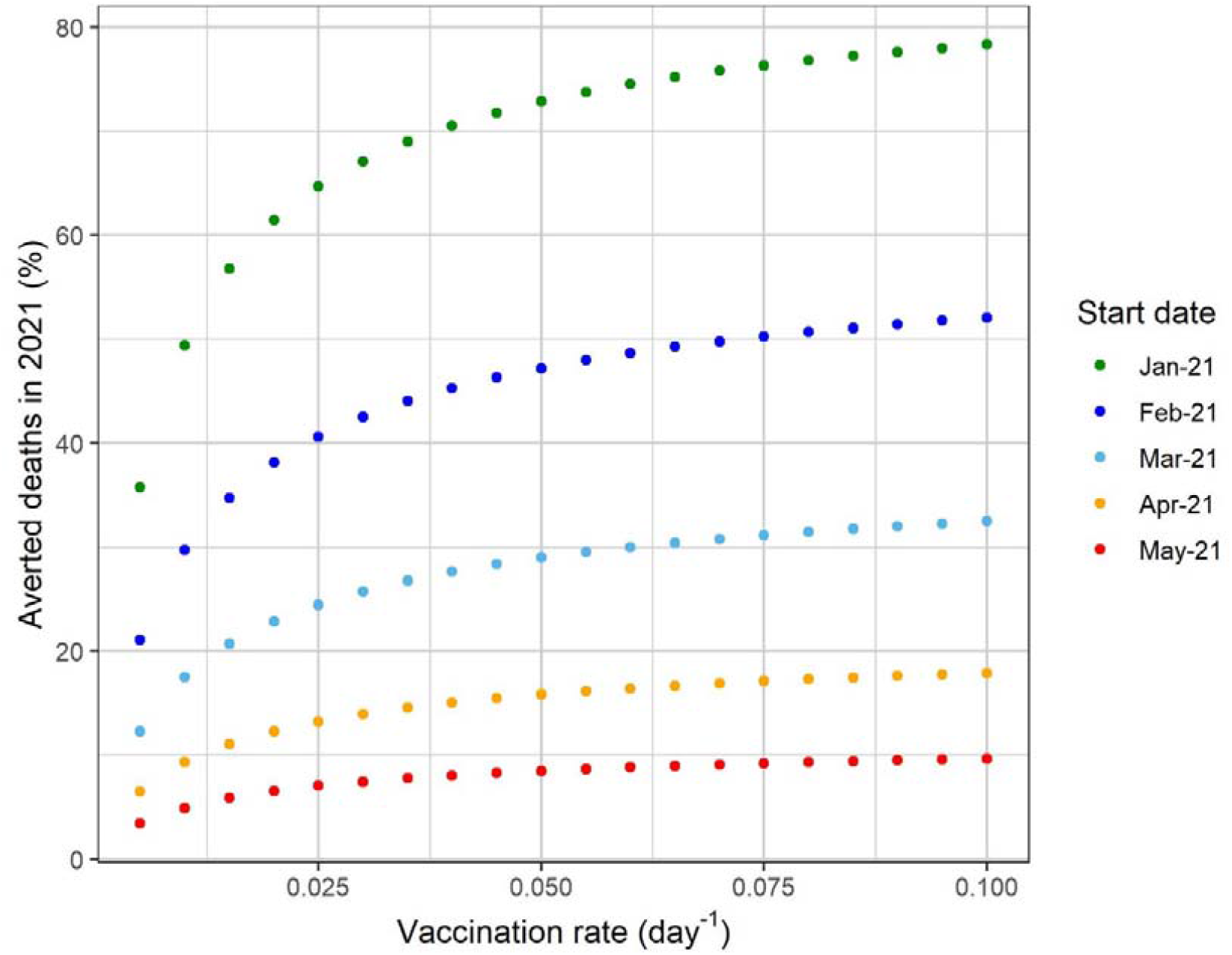
**Percentage of averted deaths in 2021 in Brazil as a function of the vaccination rate for different start dates for the vaccination campaign. The baseline scenario (vaccination adherence of 70% and vaccine efficacy of 70%) was considered.**

In figure 6 we show the results of the same simulation as in Figure 5 with several vaccination scenarios, varying compliance, efficacy and data of the starting of the campaign.

**Figure 6.**
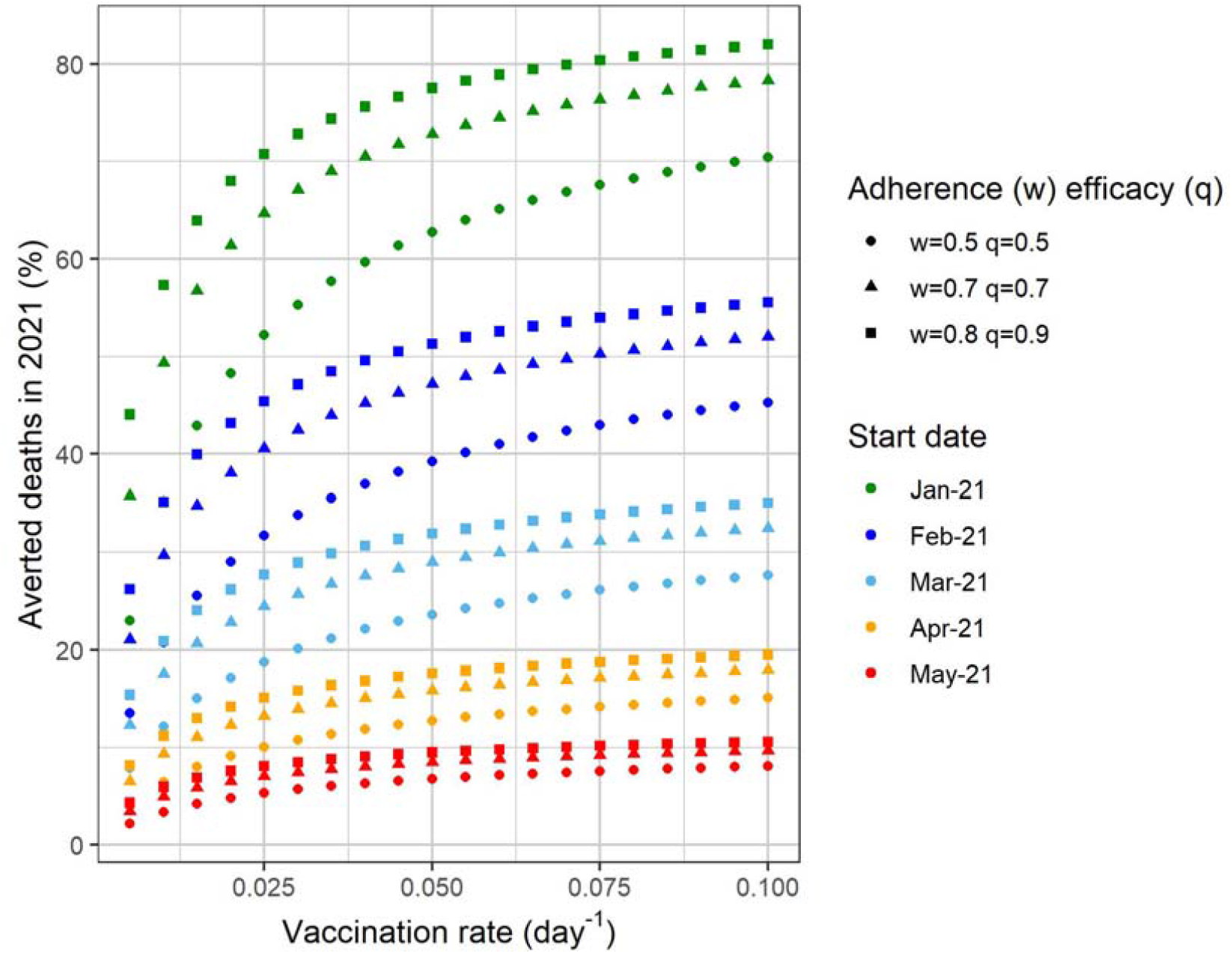
**Percentage of averted deaths in 2021 in Brazil as a function of the vaccination rate for different start dates for the vaccination campaign. Three different combinations of vaccination adherence (w) and vaccine efficacy (q) were considered: w=0.8 and q=0.9 (best-case scenario), w=0.7 and q=0.7 (baseline scenario) and w=0.5 and q=0.5 (worst-case scenario).**

In figure 7 we show the model’s projection in terms of daily new cases and deaths for an intermediate vaccination rate and for the 5 different starting dates for the campaign. In the figure, we show the simulation with a vaccination rate of 0.05 days^-1^. This implies in that with this rate the country would take approximately one year to reach the herd immunity, assumed to be 70% of the population. In addition, this vaccination rate means 544 thousand vaccinations per day, provided that there would be enough vaccine supply to this schedule.

**Figure 7.**
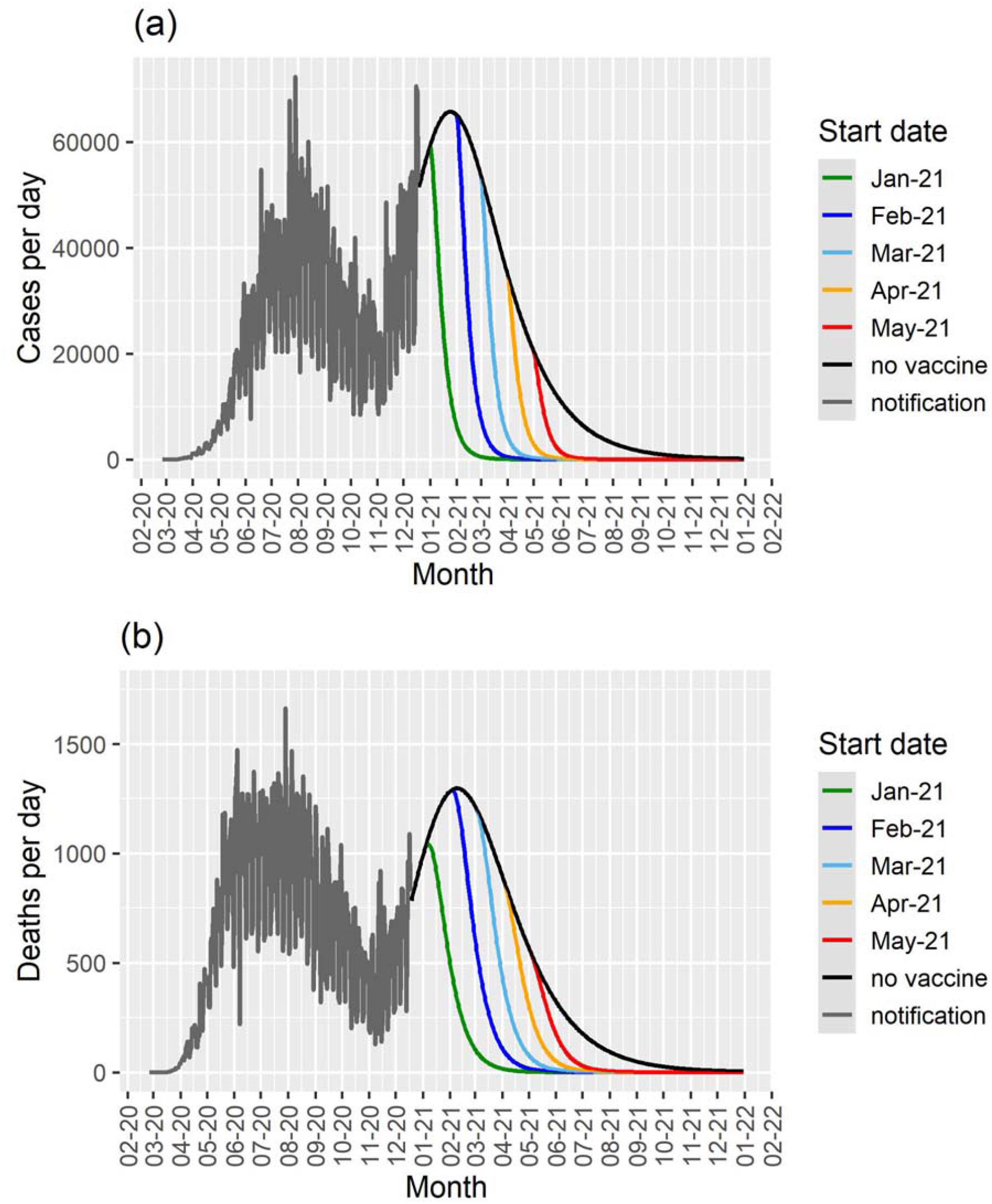
**Cases per day (a) and deaths per day (b) as a function of time for different start dates for the vaccination campaign in Brazil for a vaccination rate of 0.05 day^-1^, and the results for the model with no vaccination (black lines). The notification data until December 18^th^ 2020 are shown in gray.**

In figure 7 it is possible to observe the projected number of cases and deaths in the absence of vaccination and with the campaign beginning in January, February, March, April or May.

In figure 8 we show the same simulated scenarios as in Figure 7 with a vaccination rate ten times lower.

**Figure 8.**
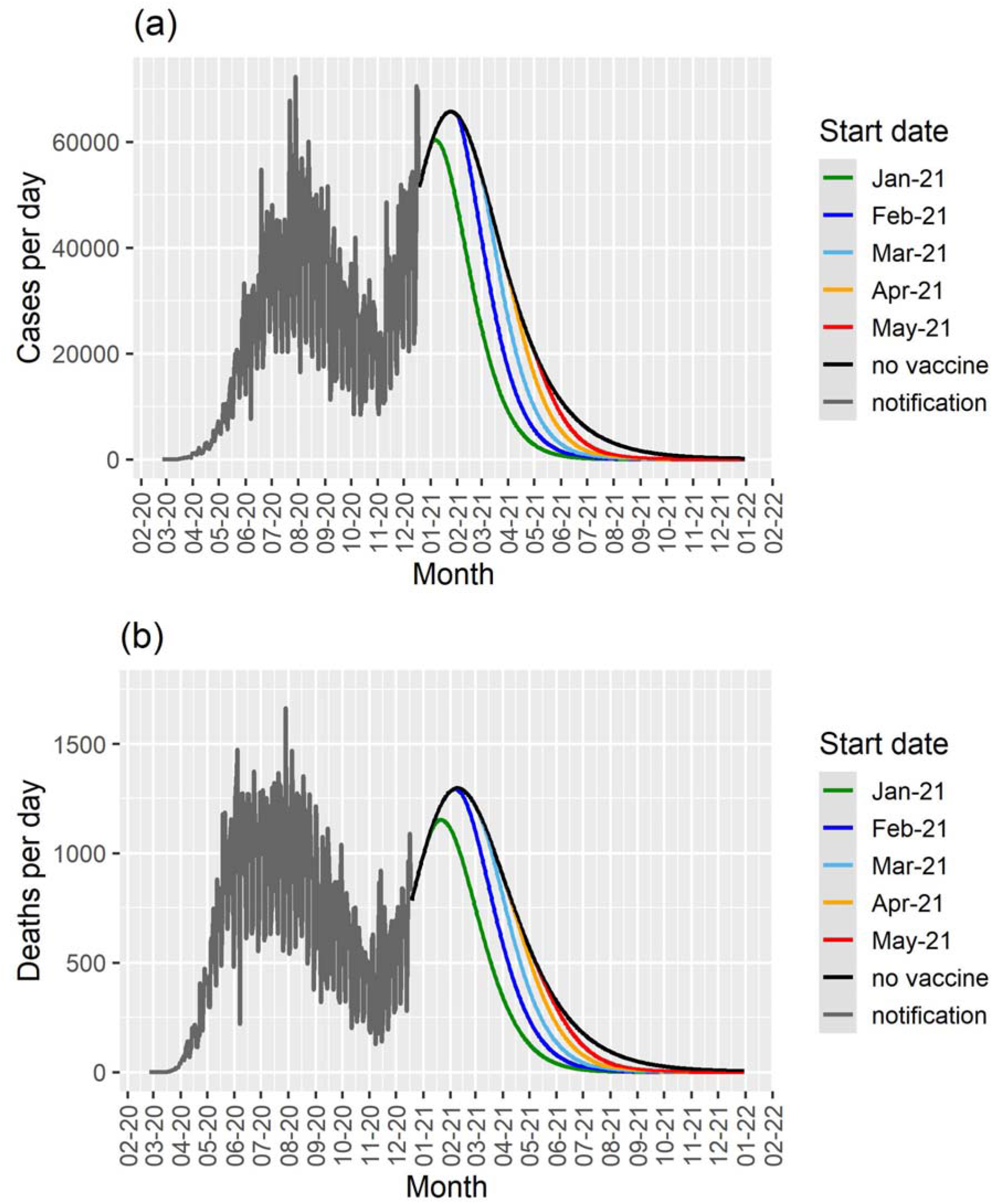
**Cases per day (a) and deaths per day (b) as a function of time for different start dates for the vaccination campaign in Brazil for a vaccination rate of 0.005 day^-1^, and the results for the model with no vaccination (black lines). The notification data until December 18^th^ 2020 are shown in gray.**

### 2) State of Sao Paulo

We fitted the model parameters simultaneously to the data of cumulative number of reported cases, deaths and the number of ICU patients (figure9) for the State of Sao Paulo until December 18, 2020. The fitting procedure is described in [16], [17].To estimate a 95% probability interval (shaded area in figure 9), we assumed a normal distribution for the contact rate with a standard deviation of 1.0%.

**Figure 9.**
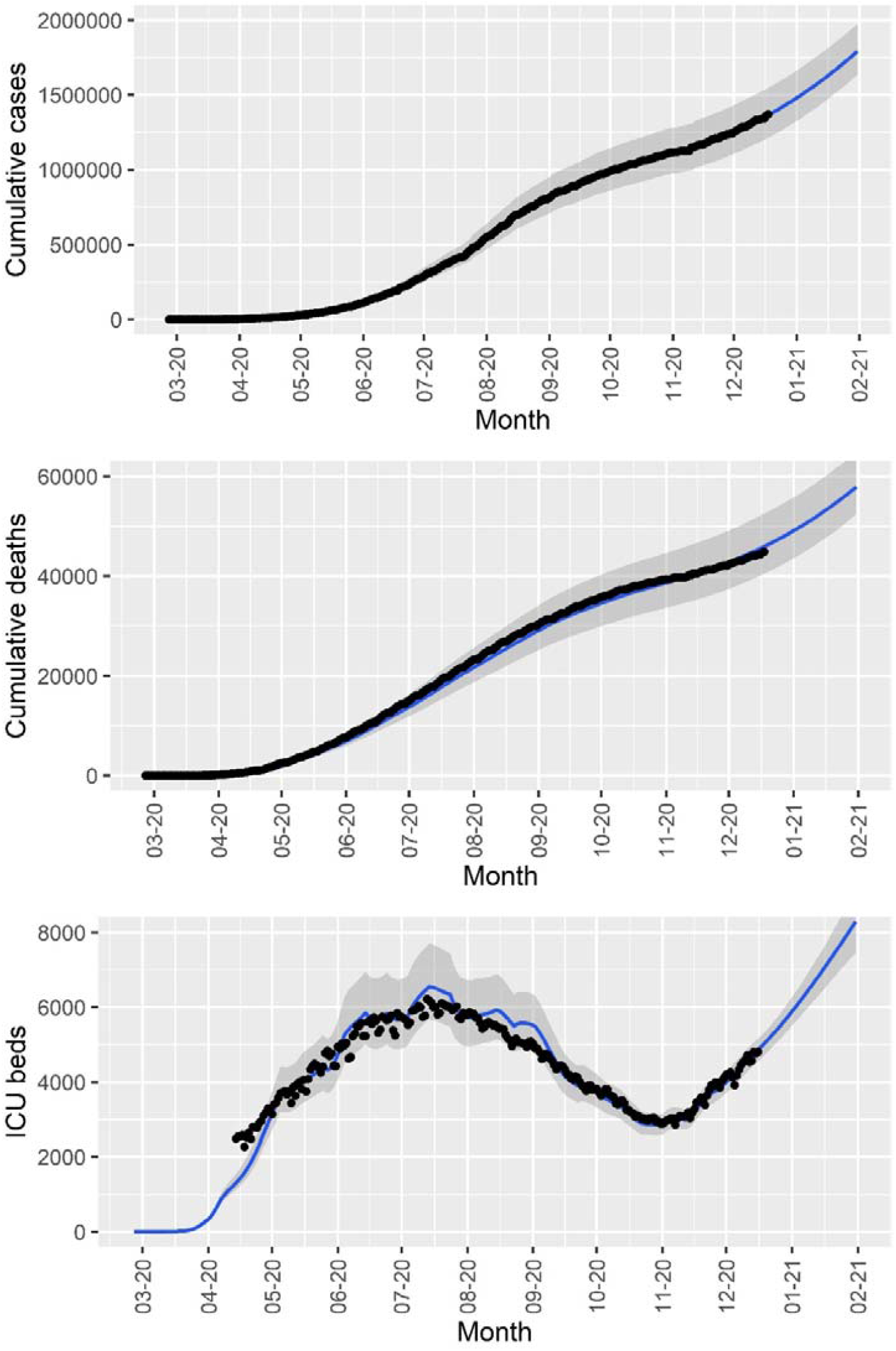
**Cumulative number of reported cases and deaths, and number of ICU patients in the State of São Paulo (black dots) and the corresponding fitted model (blue lines). The solid lines and shaded area correspond, respectively, to median values and 95% probability intervals.**

In figures 10 to 13 we show the simulations scenarios as in figures 4 to 8 for the State of São Paulo The results are qualitatively the same as in the simulation for Brazil as a whole but with magnitude proportional to the population size differences.

**Figure 10.**
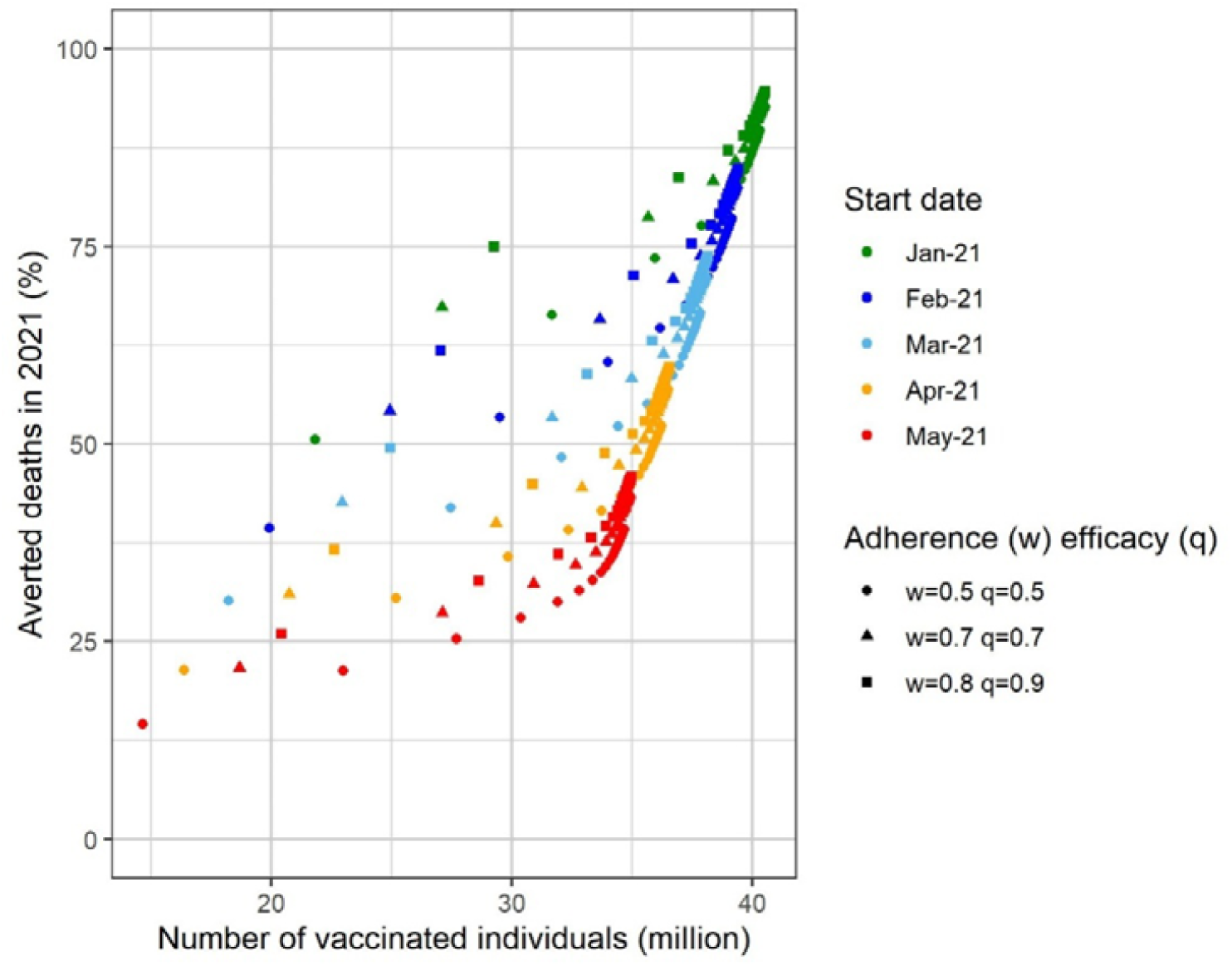
**Percentage of avoided deaths in 2021 in the State of São Paulo as a function of the number of vaccinated individuals for different start dates for the vaccination campaign. Three different combination of vaccination adherence (w) and vaccine efficacy (q) were considered: w=0.9 and q=0.9 (best-case scenario), w=0.7 and q=0.7 (baseline scenario) and w=0.5 and q=0.5 (worst-case scenario).**

**Figure 11.**
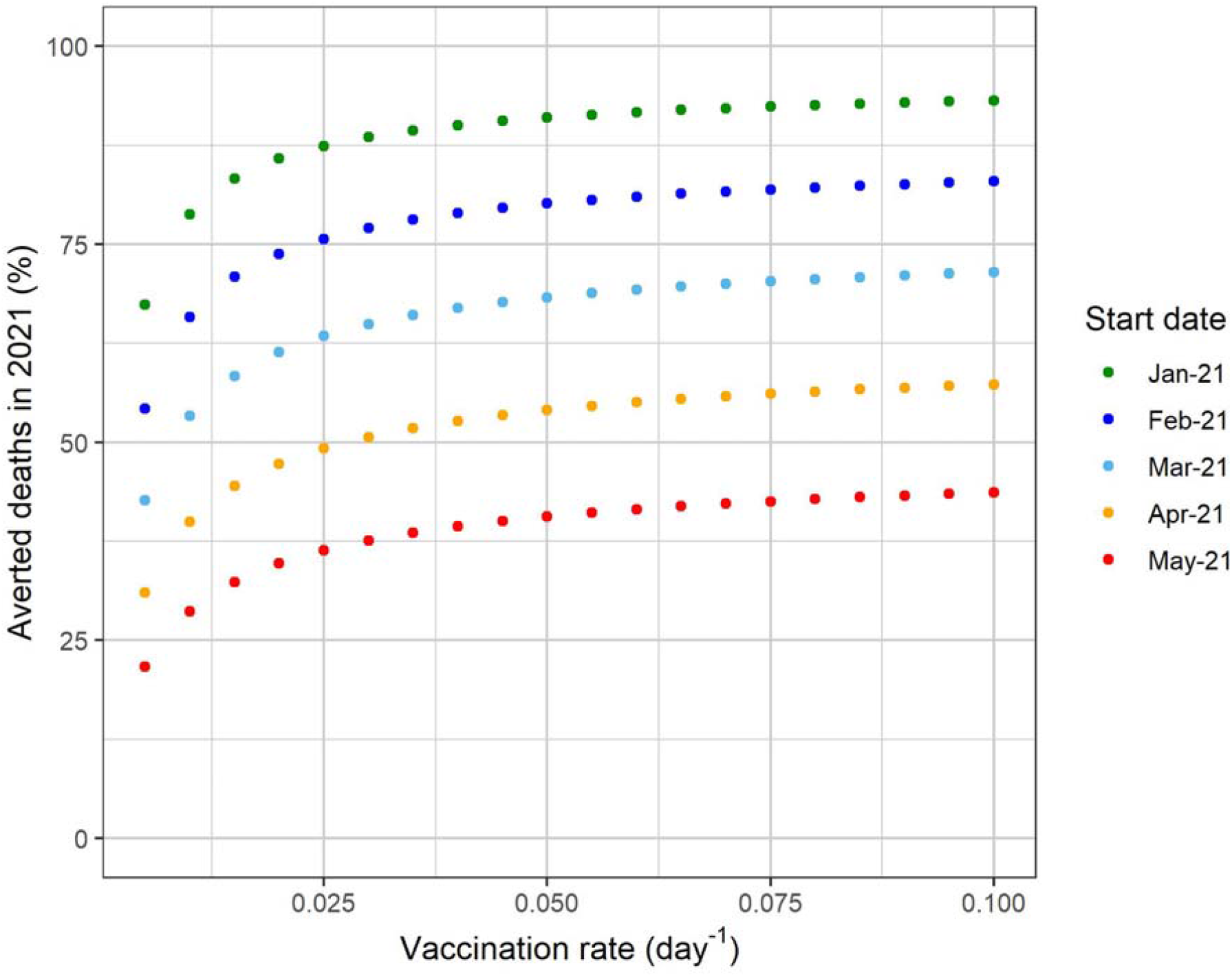
**Percentage of avoided deaths in 2021 in the State of São Paulo as a function of the vaccination rate for different start dates for the vaccination campaign. The baseline scenario (vaccination adherence of 70% and vaccine efficacy of 70%) was considered.**

**Figure 12.**
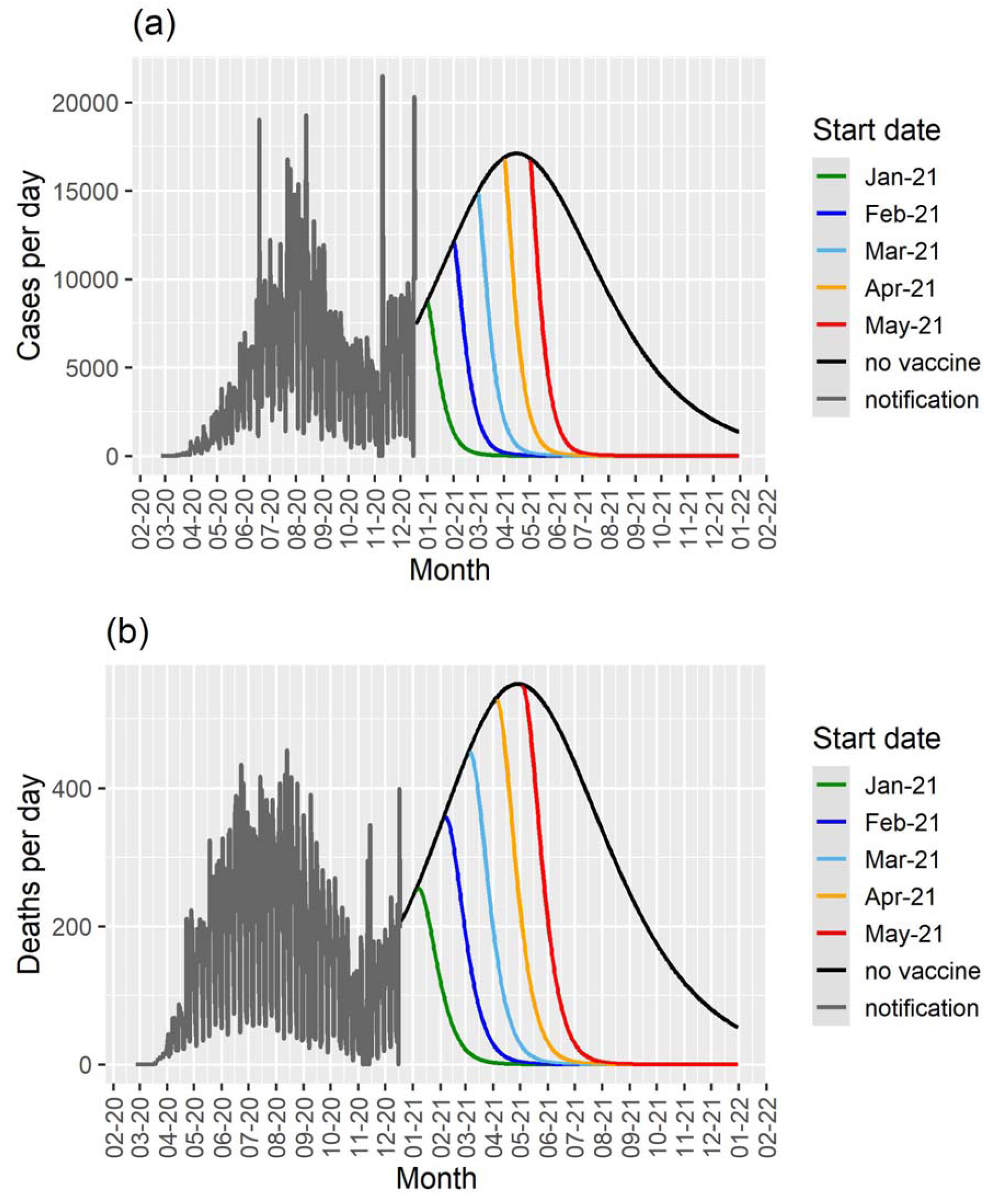
**Cases per day (a) and deaths per day (b) as a function of time for different start dates for the vaccination campaign in the State of São Paulo for a vaccination rate of 0.05 day^-1^, and the results for the model with no vaccination (black lines). The notification data until December 18^th^ 2020 are shown in gray.**

**Figure 13.**
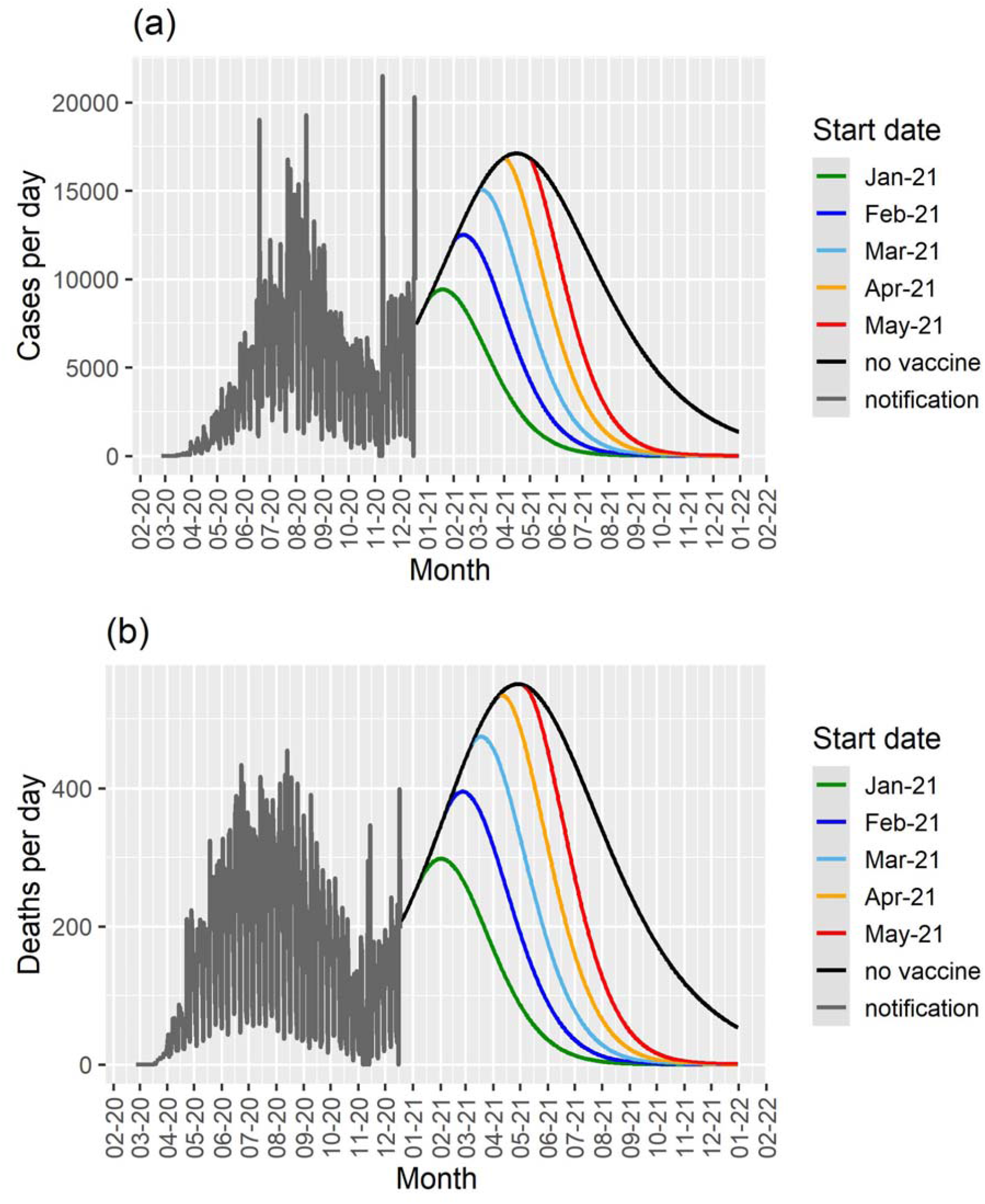
**Cases per day (a) and deaths per day (b) as a function of time for different start dates for the vaccination campaign in the State of São Paulo for a vaccination rate of 0.005 day^-1^, and the results for the model with no vaccination (black lines). The notification data until December 18^th^ 2020 are shown in gray.**

In tables 2 and 3 we summarize our main results for one particular scenario maximizing the vaccination rate (0.1 days^-1^), compliance of the population (80%) and vaccine efficacy (90%), that is, what we should expect in the optimal condition. The simulations are for Sao Paulo and Brazil as a whole.

**Table 2.** Total number of expected deaths by 31 December 2021 for an optimized vaccination

|  |  |  |
| --- | --- | --- |
|  | **Total deaths until 31 December 2021** |  |
|  | **São Paulo** | **Brazil** |
| No Vaccination | 168,290 | 352,931 |
| Beginning vaccination on 21<sup>st</sup> January 2021 | 55,489 | 225,289 |
| Beginning vaccination on 21<sup>st</sup> February 2021 | 67,118 | 266,474 |
| Beginning vaccination on 21<sup>st</sup> March 2021 | 80,384 | 298,341 |
| Beginning vaccination on 21<sup>st</sup> April 2021 | 97,093 | 322,580 |
| Beginning vaccination on 21<sup>st</sup> May 2021 | 113,545 | 336,475 |

**Table 3.** Number of additional deaths due to vaccination delay in the optimum scenario.

|  |  |  |
| --- | --- | --- |
|  | **Expected number of deaths due to vaccination delay until 31 December 2021** |  |
|  | **Sao Paulo** | **Brazil** |
| 1 month delay | 11,629 | 41,185 |
| 2 months delay | 24,895 | 73,052 |
| 3 months delay | 41,604 | 97,291 |
| 4 months delay | 58,056 | 111,186 |

It can be noted from table 2 that, in the absence of vaccination, the model projects almost 170 thousand deaths and more than 350 thousand deaths until the end of 2021 for Sao Paulo and Brazil, respectively. If in contrast, Sao Paulo and Brazil had enough vaccine supply and so started a vaccination campaign in January with the maximum vaccination rate, compliance and efficacy, they could have averted more than 112 thousand deaths and 127 thousand deaths, respectively.

In table 3 we show the number of additional deaths attributable to vaccination delay. It can be seen that for each month of delay the number of deaths increases monotonically (in a logarithm fashion) for both the State of Sao Paulo and Brazil as a whole.

## Discussion

In this paper we present a theoretical exercise, represented by a model intended to estimate the impact of (perhaps inevitable) delay in starting vaccination against SARS-CoV-2, illustrated with the epidemic situation in Brazil and in the State of Sao Paulo. The model parameters are calibrated from reports of daily COVID-19 infections, as well as published reports and it reproduces the real data with remarkable accuracy. Our results demonstrate that, both for Brazil as a whole and for the State of Sao Paulo, for each month of delaying the starting of vaccination, the number of deaths to COVID-19 is staggering high.

We assumed vaccination rates that simulates immunization of up to 70% of the whole country in 9 months, which may seem unfeasible but Brazil has a long tradition in mass vaccination campaigns [18], managing to immunize more than 20 million people in a single day [19]. Therefore, the maximum vaccination scenario would be a real possibility, given the country experience in mass vaccination schedules adopted in the past. However, due to difficulties in vaccine acquisition, the number of available doses so far has been very low indeed [21]. At the time of writing, Brazil has vaccinated slightly above 2% of its population, way below the target of at least 70% to achieve the assumed herd immunity level.

The model has some important limitations worth mentioning, the most important is perhaps that it does not consider age-dependence in incidence of the infection and in the mortality rates. However, the model was intended to simulate a mass vaccination campaign that would include all age strata in a relatively short period of time. In addition, we considered only the original variant of the virus, which means that our results represent a lower bound in the number of cases and deaths due to vaccination delay. The current scenario of the pandemic, in which new variants of SARS-CoV-2 are emerging in some countries [20] should be considered in the simulation of future vaccination models, but there is not enough empirical evidence of the impact of these new variants as related to the vaccine efficacy.

In conclusion, our model shows that the current delay in the vaccination schedules, that is observed in many countries, has serious consequences in terms of mortality by the disease and should serve as an alert to health authorities to speed the process up such that the highest number of people to be immunized is reached in the shortest period of time.

## Data Availability

The data can be requested to the author at the

https://www.seade.gov.br/coronavirus/#

## Acknowledgements

This work was partially supported by LIM01-HFMUSP, CNPq, FAPESP and Fundação Butantan.

